# Home food procurement associated with improved food security during the COVID-19 pandemic

**DOI:** 10.1101/2023.06.01.23290848

**Authors:** Meredith T. Niles, Ashley C. McCarthy, Jonathan Malacarne, Sam Bliss, Emily H. Belarmino, Jennifer Laurent, Scott C. Merrill, Sarah A. Nowak, Rachel E. Schattman

**Affiliations:** Department of Nutrition and Food Sciences, University of Vermont, Burlington, VT, USA; Gund Institute for Environment, University of Vermont, Burlington, VT, USA; School of Economics, University of Maine, Orono, ME, USA; Rubenstein School of Natural Resources, University of Vermont, Burlington, VT, USA; Department of Nursing, University of Vermont, Burlington, VT, USA; Department of Plant and Soil Sciences, University of Vermont, Burlington, VT, USA; Department of Pathology & Laboratory Medicine, University of Vermont, Burlington, VT, USA; School of Food and Agriculture, University of Maine, Orono, ME, USA

## Abstract

Home food procurement (HFP), including gardening, is associated with food security and improved health behaviors and outcomes. In the beginning of the COVID-19 pandemic, HFP increased in many high-income countries; yet little evidence has demonstrated what impact HFP had on food security. Furthermore, existing HFP studies are largely qualitative from unrepresentative samples, limiting population-level understanding of HFP engagement and impact. Using data from a representative sample of residents (n=988) in northern New England in the United States conducted in Spring/Summer 2021, we explore the relationship between HFP engagement in the first year of the pandemic and changes in food security status. We employ matching techniques to compare food security outcomes in households with observably similar demographic and social characteristics, and examine food security outcomes in three periods among households who do and do not participate in HFP. Our results show that nearly 60% of respondents engaged in at least one kind of HFP in the first year of the COVID-19 pandemic, with food insecure households being more likely to do HFP. Food insecure households (both newly and chronically food insecure) were also more likely to do HFP activities for the first time or more intensely than they had previously. Newly food insecure households were the most likely to engage in HFP overall, especially gardening. Furthermore, HFP engagement early in the COVID-19 pandemic is associated with improved food security for food insecure households in the 9-12 months after the start of the pandemic, though these improvements were primarily associated with newly, not chronically, food insecure households. Future research about HFP should continue to explore multiple HFP strategies and their potentially myriad relationships to food security, diet, and health outcomes.

## Introduction

Producing or obtaining one’s own food via gardening, fishing, foraging, hunting, raising animals, and/or preserving food (hereafter called home food procurement (HFP) may have important effects on food security and dietary intake. Most prior research has focused on gardening (both home and community), which has been shown to increase food security (1–3), increase fruit and vegetable consumption (1,4), reduce food costs (5), and provide additional income-generating opportunities (3). However, the prevalence of other HFP activities (e.g. fishing, hunting, foraging) and their impact on food security is less understood, especially outside of indigenous communities (6). Furthermore, much of the existing evidence consists of small-scale studies (e.g., (7–9)) and qualitative case studies (10), with calls for more quantitative population-level studies (11). As a result, population-level conclusions about the prevalence of HFP and its implications for food security remain limited.

The onset of the COVID-19 pandemic brought with it a documented increase in HFP and a new opportunity to explore its impact at scale. Several COVID-19-era studies have examined individuals’ increased interest in HFP across diverse socioeconomic and political regions, including Canada (12,13), Palestine (14), Sri Lanka (15) and Chile (16). In the context of population-level disruptions to work, personal, and social lives, this literature finds various motivations for the growth in HFP during the pandemic, including food security and supply chain concerns (17–19), a desire to spend time in nature (13,14,20), more free time (13), seeking spaces of refuge and community (in community gardens) (20,21), stress reduction or mental wellbeing (16,20), and a perception that HFP activities done in the outdoors were safe (22). While most pandemic-related HFP studies have focused on gardening, Clouse et al. (2022) also documented an increase in urban foraging. Additionally, previous work demonstrated increased participation, both in terms of rates and intensity, in fishing, foraging, hunting, raising backyard animals and canning during the first six months of the pandemic (6).

While research since the onset of the COVID-19 pandemic has shown an increase in HFP across disparate global regions, there remains very little evidence about the effects of this increase in HFP on individuals and communities. The most common documented impacts of HFP engagement include benefits to mental health (16,23) and higher fruit and vegetable intake among those engaging in HFP as compared to those who do not engage in HFP (6,15), though these associations have been measured at only single time points. Similarly, little is known about the extent to which HFP activities have continued beyond the early days of the pandemic, when stay at home orders and quarantine were the norm, and people may have had additional free time (13).

Barriers to undertaking and sustaining HFP are well documented, both before and during the pandemic. Previously studied barriers include inadequate land access (7,16), limited knowledge of how to engage in HFP practices (7,16,24), shortages of supplies such as seeds (20), and high levels of pests in gardening (8). Such barriers are often enough to reduce or stop HFP altogether. For example, Chenarides et al. (2021) identified a reduction in HFP between 2017 and at the beginning of the pandemic in 2020 in two urban gardens in Phoenix and Detroit US, suggesting the fragility of participation in such activities (25).

At the outset of the pandemic, our research group deployed multiple rounds of surveys in two rural US states (Vermont and Maine) to assess how COVID-19 affected food security status, mental health, dietary intake, and other measures of wellbeing. Here, we build upon our previous work (i.e., (6,23) and expand our analysis to include data from across the first year of the pandemic related to HFP engagement before and since the COVID-19 pandemic as well as food security. Quantitative survey data collected from a representative population-level survey of nearly 1,000 respondents in Vermont and Maine was used to conduct our analysis using a series of statistical tests and matching techniques. In particular, we assessed the following research questions:

1. How did food security and HFP prevalence change during the first year of the pandemic as compared to before the pandemic?
2. Did HFP engagement during the first year of the pandemic correlate with improved food security outcomes, especially for households that were food insecure during the early part of the COVID-19 pandemic?
3. How likely are respondents to continue HFP in the future? Who is most likely to intend to continue?

## Methods

### Data Collection

Data collection was conducted in Spring/Summer 2021 in Vermont and Maine, USA. The survey builds on work by the National Food Access and COVID research Team (NFACT) (26) and expands the set of questions related to HFP participation and its barriers. The original NFACT survey underwent validation in Vermont with 25 respondents aged 18 and over (27). Institutional Review Board approval was obtained from the University of Vermont (IRB protocol 000000873) before beginning data collection. Data was collected via Qualtrics (Provo, UT) research panels. We used recruitment quotas for our general population sample to ensure that the sample was representative of the populations of Vermont and Maine with respect to race and ethnicity, based on the most recent population profiles from the American Community survey (28).

### Variables of Analysis

We used four categories of variables in our analysis (Supplementary Table 1). These were: food security, home food production since the COVID-19 pandemic (HFP COVID), increased HFP since the COVID-19 pandemic (HFP More), and demographic characteristics.

Food security status was measured using the six item short-form USDA food security module (29). Following the standard protocol for calculating food insecurity, respondents who responded affirmatively to two or more questions out of six were classified as food insecure. This binary food security measure was calculated during each of three time periods: 1) pre-COVID-19 pandemic (“Pre-COVID”-i.e., in the year before the COVID-19 pandemic); 2) Early COVID-19 pandemic (“Early COVID”-i.e., in the first year of the pandemic); and 3) Later COVID-19 pandemic (“Later COVID”-i.e., in the last four months before the survey, corresponding to Winter/Spring 2021) (Figure 1). In addition, we generated a categorical variable with three categories of food insecurity: 1) never food insecure (before or during the COVID-19 pandemic), 2) newly food insecure (food secure before the pandemic, but food insecure in early COVID), and 3) chronically food insecure (food insecure both before and in early COVID).

**Figure 1.**
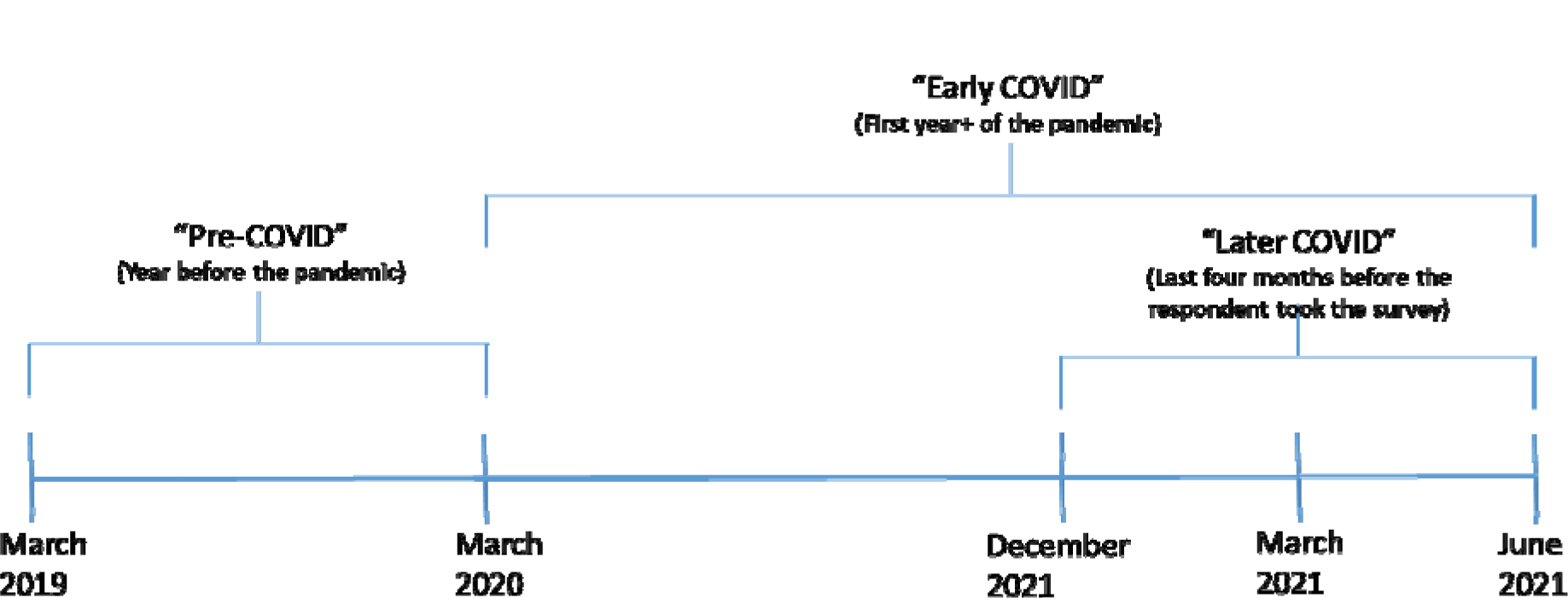
Time periods referenced for food insecurity in this article.

In addition to these key variables, we utilized several variables related to HFP, including “any HFP since March 2020” and “increased HFP since March 2020” (engaging in a HFP activity for the first time or more than before). We also explored engagement in specific HFP activities including gardening, fishing, foraging, hunting, raising livestock for meat or dairy, raising poultry for eggs, and preserving food. We inadvertently left “hunting more” out of our survey; therefore, the analyses where we explored engaging in HFP more (for the first time or more than before) include all activities except hunting.

We also report demographic characteristics of our respondents, including gender identity, race, ethnicity, income, job loss experienced during the pandemic, education, and rural/urban classification (as assessed using rural-urban commuting area (RUCA) codes) (30,31).

### Statistical Analysis and Matching Techniques

We employed several different statistical approaches to answer our research questions. Chi-Square tests were used to determine whether food insecure households were more likely to engage in HFP and analysis of variance (ANOVA) to assess whether HFP varied by food security status. We also used Kruskal Wallis tests (one-way ANOVA on ranks) to study future intention to engage in HFP.

To further explore research questions 1 and 2, we employed a quasi-experimental matching method to assess whether HFP engagement correlates with food security outcomes overall and whether HFP results in improved food security outcomes from one time period to the next. Matching analysis is a statistical approach to conditioning on observables in order to identify the effect of a “treatment” that some individuals have received and others have not (32). In our case, the treatment we are interested in is HFP engagement. We employ matching for two analyses. First, we assess whether HFP engagement correlates with food security during early and later COVID periods. Second, we analyze whether households that engaged in HFP during early COVID had improved food security status in later COVID. Our analysis matches respondents engaging in HFP to observably similar households not engaging in HFP, with an aim of balancing the distribution of both observable and unobservable covariates in each group (32). We match on a set of respondent characteristics including: race/ethnicity (BIPOC/non-Hispanic White), income (households making less than or more than $50,000 annually), gender identity (male/female^1^), job loss during the pandemic, bachelor’s degree, and rural/urban status.

We use a k-nearest neighbor matching approach, which uses the k most similar non-treated observations to create a comparison value for each treated observation. Previous research has demonstrated that this matching approach works well with eight or fewer covariates (32,33).

We report the total number of control, treated, and matched individuals in all our models to satisfy the common support condition (34). Our primary results use the Mahalanobis distance between treated and non-treated observations to identify matches and weight comparison values. Given the discrete nature of our matching variables, most of our matches are exact. In order to assess the robustness of our primary results, we repeat our estimation, varying the minimum required matches (from five down to one) and requiring that all matches be exact matches.

## Results

### Demographic Characteristics

A total of 988 individuals, including 426 in Vermont and 562 in Maine responded to the survey. Survey respondents were representative of the Vermont and Maine populations in their race/ethnicity and education. There were no major differences in outcomes by state, thus, Vermont and Maine respondents were combined for this analysis. Table 1 details the demographic characteristics of the respondents.

**Table 1.** Demographic characteristics of survey respondents and population overall of Maine

| Demographic Characteristic | Sample<br>(%) | n = 988 | U.S.<br>Population<br>(%) |
| --- | --- | --- | --- |
| Female | 68.3 | 675 | 50.9 |
| Male | 30.3 | 299 | 49.1 |
| Other gender identity/prefer not to respond | 1.4 | 14 | -- |
| Non-Hispanic White | 91.6 | 905 | 93.1 |
| BIPOC | 8.4 | 83 | 6.9 |
| Income < \$50,000 | 53.3 | 527 | 42.5 |
| Income > \$50,000 | 46.7 | 461 | 57.5 |
| Job loss during pandemic | 14.6 | 144 | -- |
| Bachelor's degree or higher | 33.9 | 335 | 33.8 |
| Rural | 55.4 | 547 | 53.5 |
| Urban | 44.6 | 441 | 46.5 |

According to their retrospective responses to the USDA food security module a year into the pandemic, 27.2% of the respondents were food insecure pre-COVID-19 (i.e., prior to March 2020), with 35.7% food insecure in early COVID, and 31.4% food insecure in later COVID (Figure 2). Among those with early food insecurity, 24.9% were chronically food insecure (pre-and early COVID), while 10.7% of respondents were newly food insecure.

**Figure 2.**
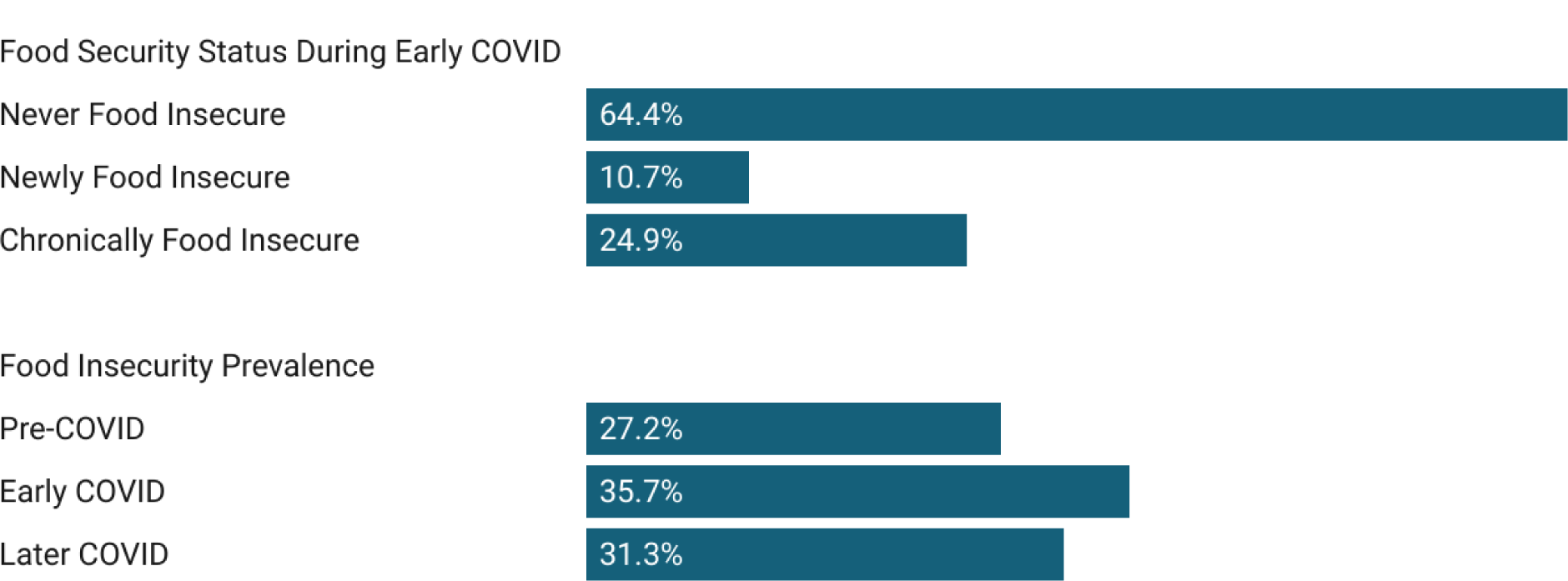
Percent of respondents classified by food security status during early COVID, and prevalence of food insecurity over time.

### Home Food Production

Nearly 60% of respondents indicated that they engaged in some type of HFP activity since March 2020, with 54.1% of those engaging in HFP indicating that they either did so for the first time or did so more since the start of the pandemic (Figure 3). Gardening was the most frequently reported HFP (46.8% of respondents), while the least frequently reported was raising livestock for meat or dairy (9.9%).

**Figure 3.**
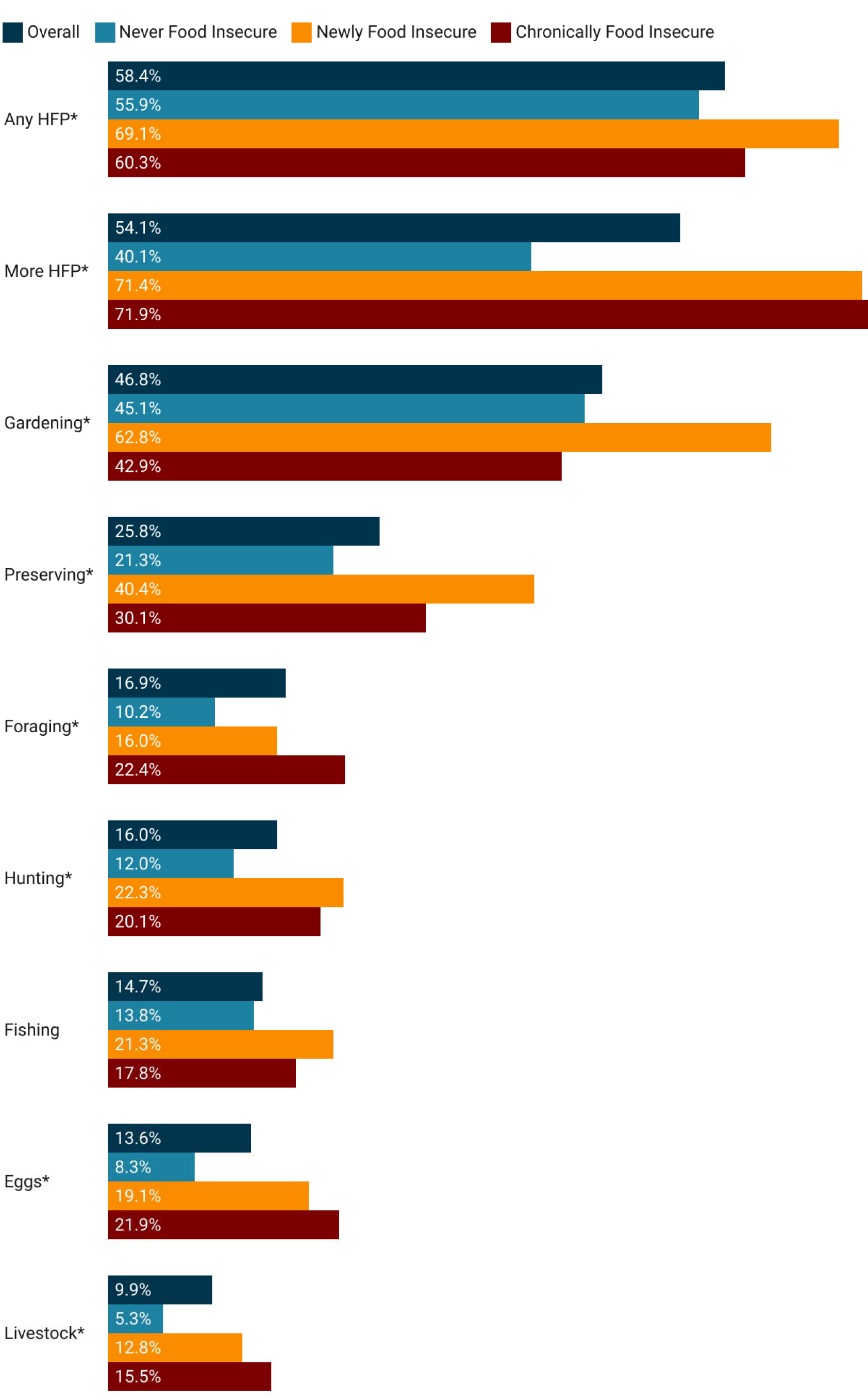
Home food production during the first year of the COVID-19 pandemic and for specific activities based on food security status. *Indicates statistically significant difference (p<0.05) across the three types of food security.

We found statistically significant differences in overall HFP engagement since the onset of the COVID-19 pandemic, as well as in specific types of HFP engagement, across food security status. Overall, food insecure households were significantly more likely to engage in HFP since the beginning of the pandemic as compared with food secure households. Newly food insecure respondents were the most likely (69.1%) to engage in HFP. Newly and chronically food insecure households were also significantly more likely than food secure households to engage in HFP for the first time or more intensely since the pandemic (p<0.001). Among specific activities, food insecure households were significantly more likely than food secure households to engage in all individual HFP activities with the exception of fishing (p=0.101). Newly food insecure households were significantly more likely to garden since the beginning of the pandemic (62.8%) but food secure and chronically food insecure households gardened at nearly the same prevalence (45.1% and 42.9% respectively).

### Changes in Food Insecurity Associated with HFP

As expected, based on the distribution of food insecure households engaging in different HFP activities, our first matching analysis (Supplementary Table 2. Robustness checks in Supplementary Table 6) identified positive associations between overall engagement in HFP and food insecurity during the first year of the pandemic. We also find that overall engagement in HFP, as well as specific HFP activities since the COVID-19 pandemic (foraging, hunting, livestock, eggs, and preserving), are positively associated with food insecurity during the first year of the pandemic. Looking at only later COVID food insecurity, any HFP engagement since the COVID-19 pandemic or gardening are not associated with food insecurity, but nearly all other activities, and engaging in them for the first time or more, continue to be positively associated with food insecurity (Supplementary Table 3. Robustness checks in Supplementary Table 7).

We find more nuanced results exploring these relationships conditional on initial food security status (Supplementary Tables 4 and 5. Robustness checks in Supplementary Tables 8 and 9). Among households that were food secure pre-COVID, those that engaged in HFP during the pandemic were more likely to be food insecure in early COVID compared to those that did not. These results were also consistent for gardening, preserving food, and those engaged in more HFP in each specific activity (Supplementary Table 4).

Among households that were food insecure pre-COVID, however, those that engaged in HFP during the pandemic were not more likely to be food insecure than those that did not engage in HFP. Taking the analysis one step further, we examined whether early COVID food insecure households who engaged in HFP at that time changed their food security status in later COVID.

Among households that experienced food insecurity early in the pandemic, those who engaged in HFP were more likely to be food secure later in the pandemic compared to those who did not engage in HFP (Supplementary Table 5). We found the same result for households that were food insecure in early COVID and gardened or foraged more than before or for the first time: they were more likely to be food secure in later COVID than those who did not. Thus, HFP engagement in early COVID is associated with improved food security outcomes for food insecure households in the 9-12 months after the onset of the pandemic.

These improvements in food security were primarily associated with newly food insecure households. Among newly food insecure households, 21.5% became food secure in later COVID, while only 9.7% chronically food insecure households became food secure (p=0.005). Furthermore, when examining these changes by HFP participation (Figure 4), we find that newly food insecure households that engaged in HFP had the highest conversion to food security in later COVID (25.0%), as compared to chronically food insecure households also engaging in HFP (10.7%,), newly food insecure not doing HFP (13.8%) and chronically food insecure not engaging in HFP (8.2%) (p=0.010).

**Figure 4.**
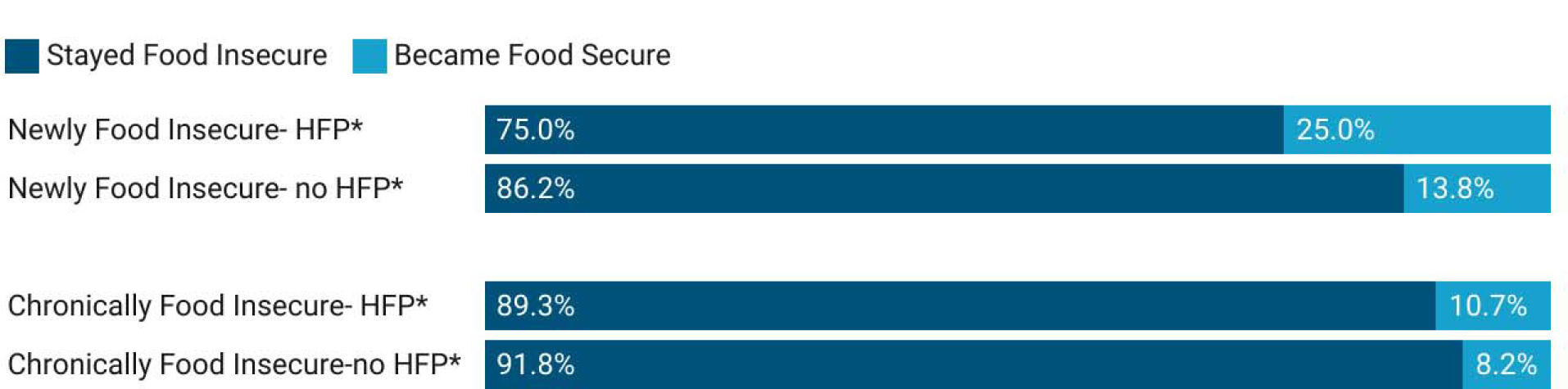
Change in food security status (in the four months prior to the survey) among newly and chronically food insecure households engaging or not in HFP. *Indicates statistically significant difference (p<0.05) between food security improvement by HFP status.

### Likelihood to Continue HFP

Finally, we examined the likelihood of respondents to continue HFP in the coming year (2021–2022). Overall, 80.7% of respondents indicated they intended to engage in some type of HFP activity in the coming year, with engagement in gardening being the most likely (70.9%) (Figure 5). Just over one-quarter of respondents indicated they would try a new HFP activity in the upcoming year (27.7%) or engage in at least one of their existing HFP activities more than previous (25.5%). Overall, both newly and chronically food insecure households were significantly (p<0.001) more likely to intend to continue all types of HFP in the coming year and would increase their engagement with HFP activities.

**Figure 5.**
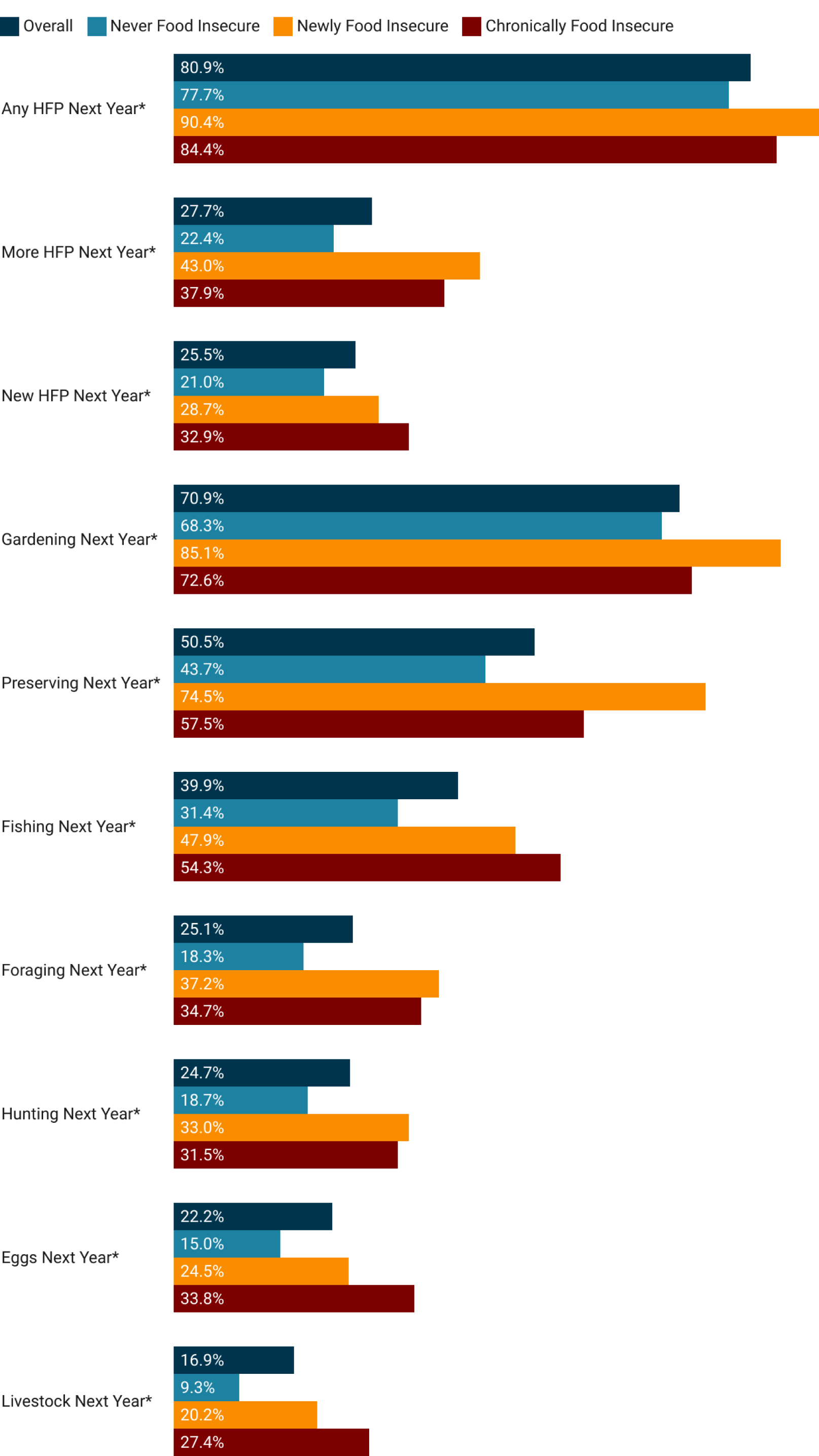
Respondent reports of intent to continue HFP activities and intensity in the 2021-2022 year. *Indicates a statistically significant difference (p<0.001) among outcomes by food security status.

## Discussion

Despite the growth in research exploring HFP during the COVID-19 pandemic, this work is among the first to directly correlate HFP during the COVID-19 pandemic with improved food security outcomes for food insecure households. Our research also demonstrates that, among this sample of respondents in two rural New England states, most households engage in HFP and engagement in these activities continued to grow during the first year of the pandemic, especially among food insecure households. This is particularly evident in comparison to previous results, which found that 35% of respondents, from a sample in the same region with similar demographic characteristics, engaged in HFP during the first six months of the COVID-19 pandemic (6). By comparison, 58% of respondents in this study engaged in some HFP activity within the first year of the pandemic.

These comparisons also point towards important future research needs related to measuring HFP at multiple time points throughout the year. For example, in the first six months of our previous analysis, only 6.2% of respondents engaged in hunting from March to August 2020 (6); but the current study shows 16.0% of respondents engaged in hunting within the first year of the pandemic. The difference may be attributed to temporal variations in hunting seasons for Vermont and Maine for game that occur in the late fall and early winter, a timepoint which the first iteration of our survey failed to capture. Similar differences are observed for foraging, where only 9.2% of individuals participated in the first six months of the pandemic (6), but 16.9% participated within the first year of the pandemic (accounting for fall foraging). Thus, as additional research on HFP continues, especially that which focuses on multiple HFP strategies, it is critical to consider the seasonality of those activities and design data collection that captures these factors.

These results add nuance to the existing research on HFP and food security by separating out households that have been chronically food insecure from those that were newly food insecure. Evidence from the first year of the pandemic demonstrates that newly versus chronically food insecure households had different demographic and social experiences with food insecurity, which may have influenced their engagement in HFP. A review of food security dynamics in the U.S.A. shows that chronic food insecurity is most likely to be experienced by non-white, less educated individuals in households headed by women (35). It is noted that it is difficult for current measures of food insecurity to fully capture the processes that lead to chronic food insecurity. At the same time, it is important to better understand the dynamic ways in which social and environmental shocks, like those experienced during the COVID-19 pandemic, influence the severity and persistence of food insecurity (36). Likewise, motivation to engage in HFP can vary based on socio-demographic factors, prior experience, available time, and other factors, which not only affect whether an individual engages in HFP, but how they do so. For example, a recent study found that women gardeners were more likely than men to plant a diversity of plant species, and that region of origin influences crop composition choices (37). Whether someone engages in HFP activities, such as hunting and fishing, as supplementary versus subsistence food acquisition behaviors can be influenced by cultural traditions, such as those held by some members of Tribal communities (38). Furthermore, a study of post-communist countries in the European Union explored how HFP varied between being largely recreational to being a coping strategy for food security, with differential impacts on well-being (39).

Our previous work identified that food insecure households were not more likely to engage in HFP overall, but instead were more likely to be engaging in HFP more or for the first time since the onset of the pandemic (6). However, this analysis finds that the story is more complicated. Newly food insecure households are the most likely to have engaged in HFP. Although, overall, chronically and newly food insecure individuals equally engaged in HFP more or for the first time, they engaged in certain, specific HFP activities at significantly different levels. Newly food insecure individuals were significantly more likely to garden and preserve food for example, while chronically food insecure households were more likely to forage. The relative influence of gardening on improving food security outcomes, especially for the newly food insecure, may be a function of the amount of time, resources, or knowledge dedicated to the activity. Future research in this area should explore the barriers to HFP by food security status, to better understand the ways to support HFP activities by diverse households.

That newly food insecure households, especially those who gardened, were more likely to transition back to food security in the first year of the pandemic continues to add evidence to the existing body of research demonstrating that gardening correlates with improved food and nutrition security (7,40–42). Our work strengthens these findings, particularly given our large sample size across representative demographics in two states, which builds upon the largely qualitative or non-representative structure of previous studies. Our work also demonstrates that improved food security associated with HFP and gardening occurs within crises, as well as in “normal” times, as evidenced by others already (e.g. (1,3).

The prevalence of HFP during a crisis period is noteworthy, and important for future research. The global COVID-19 pandemic deeply affected the global economy and food systems (43,44). As the direct financial and public health restrictions and impacts from the pandemic have waned, it remains unclear the extent to which the interest in HFP will continue, especially as individuals resume employment and additional activities. The pandemic provided many people the opportunity to engage in new activities with time commitments that may not be feasible or desirable in non-pandemic crises (13). Indeed, since our measure of food security utilizes a survey instrument that largely captures whether someone has enough money for food, these dynamics and relationships would likely change as “normal” life resumed and people regained assured access to income and groceries. At the same time, there are continued long-term impacts of the pandemic, including elevated levels of anxiety and depression (45,46), which may motivate many people to continue HFP for other mental health and well-being associated reasons (14,16,20).

Future research may, and should, continue to track HFP engagement as well as its contribution to food and nutrition security in the recovery from the pandemic. At the same time, additional quantitative research could assess how the food outputs from HFP relate to food, dietary intake, and related health outcomes. Most existing studies in the Global North have not examined how the percent of food obtained from HFP influences food and diet outcomes. Additional research in this vein could also more completely assess the varying potential impacts of HFP engagement beyond food security. Indeed, other studies have demonstrated that HFP is associated with improved mental health (23), physical activity (47,48), and social connectedness (49,50); but rarely are the suite of these potential impacts explored together. Ultimately, our work is limited in its ability to demonstrate causality because it is cross-sectional; future studies using cohort models could understand how people engage in HFP and its link to food security and other health and diet outcomes more concretely by tracking the same people and households over time.

## Conclusion

Here we show how HFP was used in different ways and intensities before and during the COVID-19 pandemic across two predominantly rural U.S. states. Our results reveal notable differences between segments that became food insecure during this period of upheaval and those that are chronically food insecure. It is important to note that this environment in general and the use of HFP is dynamic, and changes that occurred during the pandemic may reverse towards a previous “normal”, solidify as a new stable state, or become exacerbated by continued political, socio-ecological or economic issues such as inflation, recession, or a resurgence of disease. Given the emerging evidence that HFP can contribute to positive health behaviors and outcomes, our findings, and future research have public health importance, with relevance to audiences interested in human health and wellbeing, as well as the social and environmental consequences of mainstream food systems.

## Declarations

## Data Availability

The survey materials associated with this manuscript are available at Harvard Dataverse

https://dataverse.harvard.edu/dataverse/foodaccessandcoronavirus

## Acknowledgements

We’d like to thank additional members of our teams in Vermont and Maine including Farryl Bertmann and Kate Yerxa, and thank Maddie Alpaugh for her assistance with data collection.

## Authors’ contributions

MTN contributed to study design, data collection, analysis, writing, revisions, and project management. ACM contributed to study design, data collection, data cleaning and revisions. JM contributed to study design, data collection, data analysis and revisions. SB, EHB, JL, SCM, SAN, RES contributed to study design, data collection, and revisions.

## Ethics approval and consent to participate

Ethical approval was obtained through the Institutional Review Board at the Universities of Maine and Vermont, and written consent was obtained from all participants.

## Consent for publication

All authors consent and have approved this manuscript for publication

## Availability of data and material

The survey instruments for this work are publicly available at Harvard Dataverse, and the underlying data will be made publicly available upon acceptance.

## Footnotes

1 Our question related to gender identity included non-binary options; however, only a small sample indicated non-binary gender. Therefore, we utilize male/female gender identities only for our matching analysis.

