## Supplementary Materials for "Home food procurement associated with improved food security during the COVID-19 pandemic"

Supplementary Table 1. Variable Names, Questions and Scales utilized in the analysis.

| **Variable Type** | **Variable Name** | **Question(s)** | **Scale** |
| --- | --- | --- | --- |
| Food Security | Food Security- Pre- COVID | Six item USDA Food Security Module- Timeframe "Year before the COVID-19 pandemic" (Before March 2020) | 1= Food Insecure, 0= Food Secure (Answering affirmative to more than 2 of the 6 questions in the module indicates food insecure) |
|  | Food Security- Early COVID | Six item USDA Food Security Module- Timeframe "Since the COVID-19 pandemic" (During 2020) |  |
|  | Food Security- Later COVID | Six item USDA Food Security Module- Timeframe "In the last four months" (Winter/Spring 2021) |  |
|  | Chronically Food Insecure |  | 1= Food insecure before and since COVID |
|  | Newly Food Insecure |  | 1= Food secure before COVID, food insecure since COVID |
|  | Always Food Secure |  | 1= Food secure before and since COVID |
| Home Food Production Since the COVID-19 pandemic | HFP | Has your household engaged in any of these activities since the COVID-19 outbreak (March 2020)?... | 1= affirmative to any of the 7 specific activities, 0= no affirmatives |
|  | Garden Since | gardening (growing food to eat) | 1= yes, 0= no |
|  | Fishing Since | Fishing or harvesting shellfish to eat |  |
|  | Foraging Since | Foraging (harvesting mushrooms, picking wild or urban fruits or vegetables) |  |
|  | Hunting Since | Hunting or trapping to eat |  |
|  | Livestock Since | Raising animals for meat or dairy |  |
|  | Eggs Since | Keeping poultry for eggs |  |
|  | Preserving Since | Canning, fermenting, drying, smoking or otherwise preserving foods |  |
| More Home Food Production Since the COVID-19 pandemic | HFP More | Pursued any HFP activity for the first time last year or previously did the activity, but did it more last year | 1= Any of the 7 specific activities done for the first time or more last year, 0= activities not changed last year compared to previous years or done less than before |
|  | Garden More | Pursued this activity for the first time last year or previously did the activity, but did it more last year | 1= Done for the first time or more last year, 0= activities not changed last year compared to previous years or done less than before |
|  | Fishing More |  |  |
|  | Foraging More |  |  |
|  | Livestock More |  |  |
|  | Eggs More |  |  |
|  | Preserving More |  |  |
| Demographic Information | Gender identity | Which of the following best describes your gender identity? | 1 = Male, 2 = Female, 3 = Another gender identity, 4 = Prefer not to say |
|  | Race | What is your race? Check all that apply: American Indian/Alaska Native; Asian Indian; Black or African American; Chamorro; Chinese; Filipino; Japanese; Korean; Native Hawaiian; Samoan; Vietnamese; White; |  |
|  | Ethnicity | Are you of Hispanic, Latino, or Spanish origin? | 1 = no, not Hispanic, Latino, or Spanish origin, 2 = Yes, Mexican, Mexican American, Chicano, 3 = Yes, Puerto Rican, 4 = Yes, Cuban, 5 = Yes, Hispanic, Latino, or Spanish origin |
|  | BIPOC |  | 1= BIPOC and/or Hispanic; 0= non-Hispanic White |
|  | Income | Which of the following best describes your household income range in 2019 before taxes? | 1 = Less than $10,000, 2 = $10,000-$24,999, 3 = $25,000-$49,999, 4 = $50,000-$74,999, 5 = $75,000-$99,999, 6 = $100,000 or more |
|  | Job Loss | Have you or anyone in your household experienced a loss of income, reduction of hours, furlough or job loss since the COVID-19 outbreak began March 11th, 2020? | 1= yes, lost job, 0= no job loss |
|  | Education | What is the highest level of formal education that you have completed? | 1 = Some high school (no diploma), 2 = High school graduate (incl. GED), 3 = Some college (no degree), 4 = Associates degree/technical school/apprenticeship, 5 = Bachelor's degree, 6 = Postgraduate (like Master's, PhD) /professional degree (like JD) |
|  | Rural/Urban | What is your zip code? | Zip codes matched to RUCA codes: 1 = urban, 2 = large rural, 3 = small rural, 4 = isolated. Rural categorized as responding 2, 3 or 4 in RUCA code. |

Supplementary Table 2. Results from a matching analysis examining the relationship of HFP activities in the first year of the COVID-19 pandemic to food insecurity during the first year of the COVID-19 pandemic.

|  | Coefficient | AI Robust Std. Error | p= | 95% Confidence Interval | | Treated/control raw n= | Treated/control matched n= |
| --- | --- | --- | --- | --- | --- | --- | --- |
| HFP Since COVID | 0.077 | 0.030 | 0.010 | 0.018 | 0.136 | 527/379 | 527/527 |
| Garden Since | 0.059 | 0.030 | 0.051 | -0.003 | 0.118 | 411/472 | 411/411 |
| Fishing Since | 0.090 | 0.047 | 0.058 | -0.003 | 0.183 | 140/743 | 140/140 |
| Foraging Since | 0.172 | 0.049 | 0.000 | 0.076 | 0.267 | 121/762 | 121/121 |
| Hunting Since | 0.014 | 0.047 | 0.003 | 0.048 | 0.234 | 132/751 | 132/132 |
| Livestock Since | 0.189 | 0.062 | 0.003 | 0.065 | 0.312 | 76/807 | 76/76 |
| Eggs Since | 0.192 | 0.051 | 0.000 | 0.093 | 0.292 | 112/771 | 112/112 |
| Preserving Since | 0.127 | 0.039 | 0.001 | 0.051 | 0.204 | 220/663 | 220/220 |
| HFP More | 0.239 | 0.046 | 0.000 | 0.148 | 0.330 | 272/241 | 272/272 |
| Gardens More | 0.176 | 0.048 | 0.000 | 0.082 | 0.270 | 186/224 | 186/186 |
| Fishing More | 0.405 | 0.085 | 0.000 | 0.238 | 0.571 | 52/88 | 52/52 |
| Foraging More | 0.342 | 0.091 | 0.000 | 0.164 | 0.520 | 61/60 | 61/61 |
| Livestock More | 0.247 | 0.123 | 0.045 | 0.006 | 0.488 | 37/38 | 37/37 |
| Eggs More | 0.282 | 0.100 | 0.005 | 0.087 | 0.477 | 62/49 | 62/62 |
| Preserving More | 0.287 | 0.066 | 0.000 | 0.156 | 0.416 | 113/107 | 113/113 |

Supplementary Table 3. Results from a matching analysis examining the relationship of HFP activities in the first year of the COVID-19 pandemic to food insecurity in the last four months before the survey (Winter/Spring 2021).

|  | Coefficient | AI Robust Std. Error | p= | 95% Confidence Interval | | Treated/control raw n= | Treated/control matched n= |
| --- | --- | --- | --- | --- | --- | --- | --- |
| HFP Since COVID | 0.040 | 0.030 | 0.179 | -0.018 | 0.098 | 509/370 | 509/509 |
| Garden Since | 0.023 | 0.030 | 0.441 | -0.036 | 0.082 | 400/461 | 400/400 |
| Fishing Since | 0.110 | 0.047 | 0.018 | 0.018 | 0.201 | 136/725 | 136/136 |
| Foraging Since | 0.175 | 0.049 | 0.000 | 0.079 | 0.271 | 117/744 | 117/117 |
| Hunting Since | 0.136 | 0.047 | 0.004 | 0.043 | 0.228 | 125/736 | 125/125 |
| Livestock Since | 0.203 | 0.065 | 0.002 | 0.075 | 0.331 | 70/791 | 70/70 |
| Eggs Since | 0.189 | 0.052 | 0.000 | 0.087 | 0.291 | 106/755 | 106/106 |
| Preserving Since | 0.110 | 0.038 | 0.004 | 0.034 | 0.186 | 214/647 | 214/214 |
| HFP More | 0.228 | 0.047 | 0.000 | 0.137 | 0.320 | 264/234 | 264/264 |
| Gardens More | 0.185 | 0.045 | 0.000 | 0.095 | 0.275 | 184/216 | 184/184 |
| Fishing More | 0.414 | 0.087 | 0.000 | 0.243 | 0.585 | 50/86 | 50/50 |
| Foraging More | 0.315 | 0.093 | 0.001 | 0.133 | 0.497 | 58/59 | 58/58 |
| Livestock More | 0.203 | 0.126 | 0.107 | -0.043 | 0.450 | 33/37 | 33/33 |
| Eggs More | 0.171 | 0.106 | 0.106 | -0.036 | 0.378 | 59/47 | 59/59 |
| Preserving More | 0.230 | 0.068 | 0.001 | 0.097 | 0.364 | 109/105 | 109/109 |

Supplementary Table 4. Results from a matching analysis examining the relationship of HFP activities in the first year of the COVID-19 pandemic to food insecurity in 2020 during the pandemic, depending on the food security status of individuals prior to the pandemic.

| HFP Activity | Food security status Year Before COVID | Coefficient | AI Robust Std. Error | p= | 95% Confidence Interval | | Treated/ control raw n= | Treated/ control matched n= |
| --- | --- | --- | --- | --- | --- | --- | --- | --- |
| HFP Since COVID | Food Secure | 0.054 | 0.027 | 0.042 | 0.002 | 0.107 | 527/379 | 527/527 |
|  | Food Insecure | 0.020 | 0.031 | 0.524 | -0.042 | 0.082 |  |  |
| Garden Since | Food Secure | 0.076 | 0.027 | 0.005 | 0.023 | 0.130 | 411/472 | 411/411 |
|  | Food Insecure | 0.018 | 0.031 | 0.576 | -0.079 | 0.044 |  |  |
| Fishing Since | Food Secure | 0.065 | 0.045 | 0.154 | -0.024 | 0.154 | 140/743 | 140/140 |
|  | Food Insecure | -0.039 | 0.047 | 0.402 | -0.132 | 0.053 |  |  |
| Foraging Since | Food Secure | 0.078 | 0.049 | 0.111 | -0.018 | 0.174 | 121/762 | 121/121 |
|  | Food Insecure | -0.001 | 0.039 | 0.973 | -0.078 | 0.075 |  |  |
| Hunting Since | Food Secure | 0.087 | 0.048 | 0.077 | -0.009 | 0.183 | 132/751 | 132/132 |
|  | Food Insecure | -0.007 | 0.038 | 0.852 | -0.083 | 0.069 |  |  |
| Livestock Since | Food Secure | 0.108 | 0.074 | 0.148 | -0.038 | 0.254 | 76/807 | 76/76 |
|  | Food Insecure | -0.029 | 0.059 | 0.626 | -0.146 | 0.088 |  |  |
| Eggs Since | Food Secure | 0.109 | 0.058 | 0.060 | -0.005 | 0.223 | 112/771 | 112/112 |
|  | Food Insecure | -0.019 | 0.041 | 0.649 | -0.100 | 0.063 |  |  |
| Preserving Since | Food Secure | 0.087 | 0.039 | 0.028 | 0.009 | 0.163 | 220/663 | 220/220 |
|  | Food Insecure | 0.031 | 0.032 | 0.327 | -0.031 | 0.095 |  |  |
| HFPMore | Food Secure | 0.162 | 0.043 | 0.000 | 0.077 | 0.246 | 260/239 | 260/260 |
|  | Food Insecure | -0.047 | 0.037 | 0.207 | -0.121 | 0.026 |  |  |
| Garden More | Food Secure | 0.167 | 0.046 | 0.000 | 0.076 | 0.259 | 186/224 | 186/186 |
|  | Food Insecure | 0.021 | 0.057 | 0.710 | -0.091 | 0.133 |  |  |
| Fishing More | Food Secure | 0.315 | 0.112 | 0.005 | 0.096 | 0.535 | 52/88 | 52/52 |
|  | Food Insecure | 0.141 | 0.135 | 0.292 | -0.122 | 0.406 |  |  |
| Foraging More | Food Secure | 0.396 | 0.101 | 0.000 | 0.197 | 0.595 | 61/60 | 61/61 |
|  | Food Insecure | -0.091 | 0.050 | 0.069 | -0.189 | 0.007 |  |  |
| Livestock More | Food Secure | 0.413 | 0.133 | 0.002 | 0.152 | 0.674 | 37/38 | 37/37 |
|  | Food Insecure | -0.037 | 0.125 | 0.763 | -0.282 | 0.207 |  |  |
| Eggs More | Food Secure | 0.305 | 0.101 | 0.003 | 0.107 | 0.503 | 62/49 | 62/62 |
|  | Food Insecure | 0.142 | 0.121 | 0.240 | -0.095 | 0.38 |  |  |
| Preserving More | Food Secure | 0.157 | 0.072 | 0.029 | 0.016 | 0.296 | 113/107 | 113/113 |
|  | Food Insecure | 0.033 | 0.081 | 0.068 | -0.125 | 0.192 |  |  |

Supplementary Table 5. Results from a matching analysis examining the relationship of HFP activities in the first year of the COVID-19 pandemic to food insecurity in the last four months (Late COVID), depending on the food security status of individuals early in the pandemic (Early COVID).

|  | Food Security Status in Early COVID | Coefficient | AI Robust Std. Error | p= | 95% Confidence Interval | | Treated/ control raw n= | Treated/ control matched n= |
| --- | --- | --- | --- | --- | --- | --- | --- | --- |
| HFP Since COVID | Food Secure | -0.001 | 0.012 | 0.967 | -0.025 | 0.024 | 509/370 | 509/509 |
|  | Food Insecure | -0.079 | 0.039 | 0.040 | -0.154 | -0.004 |  |  |
| Garden Since | Food Secure | -0.002 | 0.010 | 0.798 | -0.022 | 0.017 | 400/461 | 400/400 |
|  | Food Insecure | -0.110 | 0.042 | 0.008 | -0.192 | -0.028 |  |  |
| Fishing Since | Food Secure | 0.008 | 0.019 | 0.679 | -0.029 | 0.045 | 136/725 | 136/136 |
|  | Food Insecure | 0.033 | 0.050 | 0.507 | -0.065 | 0.132 |  |  |
| Foraging Since | Food Secure | 0.001 | 0.020 | 0.980 | -0.039 | 0.040 | 117/744 | 117/117 |
|  | Food Insecure | 0.052 | 0.055 | 0.337 | -0.054 | 0.160 |  |  |
| Hunting Since | Food Secure | 0.035 | 0.026 | 0.167 | -0.048 | 0.085 | 125/736 | 125/125 |
|  | Food Insecure | -0.011 | 0.056 | 0.842 | -0.121 | 0.099 |  |  |
| Livestock Since | Food Secure | 0.046 | 0.048 | 0.339 | -0.048 | 0.139 | 70/791 | 70/791 |
|  | Food Insecure | 0.013 | 0.056 | 0.812 | -0.097 | 0.123 |  |  |
| Eggs Since | Food Secure | 0.024 | 0.031 | 0.443 | -0.037 | 0.085 | 106/755 | 106/106 |
|  | Food Insecure | 0.003 | 0.049 | 0.948 | -0.093 | 0.100 |  |  |
| Preserving Since | Food Secure | -0.003 | 0.016 | 0.827 | -0.034 | 0.027 | 214/647 | 214/214 |
|  | Food Insecure | 0.001 | 0.043 | 0.975 | -0.084 | 0.087 |  |  |
| HFPMore | Food Secure | 0.008 | 0.012 | 0.501 | -0.015 | 0.031 | 253/232 | 253/253 |
|  | Food Insecure | 0.020 | 0.063 | 0.748 | -0.104 | 0.145 |  |  |
| Gardens More | Food Secure | 0.015 | 0.015 | 0.334 | -0.015 | 0.045 | 184/216 | 184/184 |
|  | Food Insecure | 0.018 | 0.063 | 0.773 | -0.106 | 0.143 |  |  |
| Fishing More | Food Secure | 0.021 | 0.074 | 0.771 | -0.123 | 0.166 | 50/86 | 50/50 |
|  | Food Insecure | -0.023 | 0.068 | 0.738 | -0.156 | 0.111 |  |  |
| Foraging More | Food Secure | 0.063 | 0.061 | 0.302 | -0.056 | 0.181 | 58/59 | 58/58 |
|  | Food Insecure | -0.133 | 0.055 | 0.016 | -0.241 | -0.025 |  |  |
| Livestock More | Food Secure | 0.015 | 0.130 | 0.909 | -0.240 | 0.270 | 33/37 | 33/33 |
|  | Food Insecure | -0.045 | 0.083 | 0.592 | -0.208 | 0.119 |  |  |
| Eggs More | Food Secure | -0.108 | 0.058 | 0.063 | -0.223 | 0.005 | 59/47 | 59/59 |
|  | Food Insecure | -0.041 | 0.077 | 0.601 | -0.193 | 0.111 |  |  |
| Preserving More | Food Secure | 0.002 | 0.029 | 0.948 | -0.055 | 0.059 | 109/105 | 109/105 |
|  | Food Insecure | -0.032 | 0.065 | 0.617 | -0.160 | 0.095 |  |  |

Supplementary Table 6 – Robustness Checks for a matching analysis examining the relationship of HFP activities in the first year of the COVID-19 pandemic to food insecurity during the first year of the COVID-19 pandemic (Original estimates reported in Supplementary Table 2).

|  | Min  Matches | Exact Matches | Coefficient | AI Robust Std. Error | p= | Treated/control raw n= | Treated/control matched n= |
| --- | --- | --- | --- | --- | --- | --- | --- |
| HFP Since COVID | 5 | No | 0.077 | 0.030 | 0.010 | 527/379 | 527/527 |
|  | 4 | No | 0.079 | 0.030 | 0.008 | 527/379 | 527/527 |
|  | 3 | No | 0.071 | 0.030 | 0.019 | 527/379 | 527/527 |
|  | 2 | No | 0.069 | 0.031 | 0.027 | 527/379 | 527/527 |
|  | 1 | No | 0.088 | 0.031 | 0.005 | 527/379 | 527/527 |
|  | 5 | Yes | 0.057 | 0.032 | 0.072 | 448/327 | 448/448 |
|  | 4 | Yes | 0.057 | 0.032 | 0.072 | 448/327 | 448/448 |
|  | 3 | Yes | 0.051 | 0.032 | 0.107 | 464/341 | 464/464 |
|  | 2 | Yes | 0.070 | 0.032 | 0.031 | 485/356 | 485/485 |
|  | 1 | Yes | 0.076 | 0.032 | 0.019 | 512/379 | 512/512 |
| HFP More | 5 | No | 0.239 | 0.046 | 0.000 | 272/241 | 272/272 |
|  | 4 | No | 0.239 | 0.045 | 0.000 | 272/241 | 272/272 |
|  | 3 | No | 0.251 | 0.045 | 0.000 | 272/241 | 272/272 |
|  | 2 | No | 0.265 | 0.046 | 0.000 | 272/241 | 272/272 |
|  | 1 | No | 0.268 | 0.046 | 0.000 | 272/241 | 272/272 |
|  | 5 | Yes | 0.232 | 0.051 | 0.000 | 171/180 | 171/171 |
|  | 4 | Yes | 0.231 | 0.049 | 0.000 | 181/189 | 181/181 |
|  | 3 | Yes | 0.251 | 0.049 | 0.000 | 221/216 | 221/221 |
|  | 2 | Yes | 0.254 | 0.048 | 0.000 | 231/241 | 231/231 |
|  | 1 | Yes | 0.264 | 0.048 | 0.000 | 248/241 | 248/248 |

Supplementary Table 7 – Robustness Checks for matching analysis examining the relationship of HFP activities in the first year of the COVID-19 pandemic to food insecurity in the last four months before the survey (Winter/Spring 2021) (Original estimates reported in Supplementary Table 3).

|  | Min Matches | Exact Matches | Coefficient | AI Robust Std. Error | p= | Treated/control raw n= | Treated/control matched n= |
| --- | --- | --- | --- | --- | --- | --- | --- |
| HFP Since COVID | 5 | No | 0.040 | 0.030 | 0.179 | 509/370 | 509/509 |
|  | 4 | No | 0.048 | 0.030 | 0.107 | 517/372 | 517/517 |
|  | 3 | No | 0.048 | 0.030 | 0.108 | 517/372 | 517/517 |
|  | 2 | No | 0.039 | 0.031 | 0.205 | 517/372 | 517/517 |
|  | 1 | No | 0.058 | 0.031 | 0.061 | 517/372 | 517/517 |
|  | 5 | Yes | 0.034 | 0.032 | 0.280 | 443/321 | 443/443 |
|  | 4 | Yes | 0.034 | 0.032 | 0.280 | 443/321 | 443/443 |
|  | 3 | Yes | 0.033 | 0.031 | 0.287 | 446/326 | 446/446 |
|  | 2 | Yes | 0.042 | 0.032 | 0.193 | 447/372 | 447/447 |
|  | 1 | Yes | 0.048 | 0.032 | 0.139 | 503/372 | 503/503 |
| HFP More | 5 | No | 0.228 | 0.047 | 0.000 | 264/234 | 264/264 |
|  | 4 | No | 0.240 | 0.045 | 0.000 | 270/236 | 270/270 |
|  | 3 | No | 0.253 | 0.044 | 0.000 | 270/236 | 270/270 |
|  | 2 | No | 0.255 | 0.045 | 0.000 | 270/236 | 270/270 |
|  | 1 | No | 0.257 | 0.046 | 0.000 | 270/236 | 270/270 |
|  | 5 | Yes | 0.240 | 0.049 | 0.000 | 171/177 | 171/171 |
|  | 4 | Yes | 0.242 | 0.047 | 0.000 | 185/201 | 185/185 |
|  | 3 | Yes | 0.247 | 0.048 | 0.000 | 222/213 | 222/222 |
|  | 2 | Yes | 0.258 | 0.047 | 0.000 | 231/236 | 231/231 |
|  | 1 | Yes | 0.258 | 0.047 | 0.000 | 246/236 | 246/246 |

Supplementary Table 8 – Robustness checks for matching analysis examining the relationship of HFP activities in the first year of the COVID-19 pandemic to food insecurity in 2020 during the pandemic, depending on the food security status of individuals prior to the pandemic. (Original estimates reported in Supplementary Table 4)

| HFP Activity | Food security status Year Before COVID | Min Matches | Exact | Coefficient | AI Robust Std. Error | p= | Treated/ control raw n= | Treated/ control matched n= |
| --- | --- | --- | --- | --- | --- | --- | --- | --- |
| HFP Since COVID | Food Secure | 5 | No | 0.054 | 0.027 | 0.042 | 376/276 | 376/376 |
|  | Food Insecure | 5 | No | 0.020 | 0.031 | 0.524 | 136/95 | 136/136 |
|  | Food Secure | 1 | No | 0.063 | 0.029 | 0.030 | 376/276 | 376/376 |
|  | Food Insecure | 1 | No | 0.041 | 0.039 | 0.292 | 136/95 | 136/136 |
|  | Food Secure | 5 | Yes | 0.044 | 0.027 | 0.106 | 302/225 | 302/302 |
|  | Food Insecure | 5 | Yes | 0.049 | 0.050 | 0.318 | 72/56 | 72/72 |
|  | Food Secure | 1 | Yes | 0.051 | 0.029 | 0.079 | 359/263 | 359/359 |
|  | Food Insecure | 1 | Yes | 0.022 | 0.035 | 0.529 | 116/95 | 116/116 |
| HFPMore | Food Secure | 5 | No | 0.161 | 0.043 | 0.000 | 165/202 | 165/165 |
|  | Food Insecure | 5 | No | -0.047 | 0.037 | 0.207 | 95/37 | 95/95 |
|  | Food Secure | 1 | No | 0.176 | 0.045 | 0.000 | 165/202 | 165/165 |
|  | Food Insecure | 1 | No | -0.021 | 0.048 | 0.662 | 95/37 | 95/95 |
|  | Food Secure | 5 | Yes | 0.083 | 0.047 | 0.077 | 94/137 | 94/94 |
|  | Food Insecure | 5 | Yes | -0.077 | 0.052 | 0.141 | 26/14 | 26/26 |
|  | Food Secure | 1 | Yes | 0.159 | 0.047 | 0.001 | 147/193 | 193/193 |
|  | Food Insecure | 1 | Yes | -0.033 | 0.065 | 0.609 | 60/37 | 60/60 |

Supplementary Table 9 – Robustness checks for matching analysis examining the relationship of HFP activities in the first year of the COVID-19 pandemic to food insecurity in the last four months, depending on the food security status of individuals earlier in the pandemic. (Original estimates reported in Supplementary Table 5)

|  |  |  |  | Coefficient | AI Robust Std. Error | p= | Treated/ control raw n= | Treated/ control matched n= |
| --- | --- | --- | --- | --- | --- | --- | --- | --- |
| HFP Since COVID | Food Secure | 5 | No | -0.0005 | 0.012 | 0.967 | 320/250 | 320/320 |
|  | Food Insecure | 5 | No | -0.079 | 0.039 | 0.040 | 189/115 | 189/189 |
|  | Food Secure | 1 | No | -0.0005 | 0.012 | 0.967 | 320/255 | 320/320 |
|  | Food Insecure | 1 | No | -0.071 | 0.042 | 0.092 | 189/115 | 189/189 |
|  | Food Secure | 5 | Yes | -0.004 | 0.014 | 0.759 | 269/213 | 269/269 |
|  | Food Insecure | 5 | Yes | -0.058 | 0.048 | 0.233 | 98/66 | 98/98 |
|  | Food Secure | 1 | Yes | -0.0005 | 0.012 | 0.964 | 307/246 | 307/246 |
|  | Food Insecure | 1 | Yes | 0.062 | 0.045 | 0.170 | 161/115 | 161/161 |
| HFPMore | Food Secure | 5 | No | 0.008 | 0.012 | 0.501 | 128/105 | 128/128 |
|  | Food Insecure | 5 | No | 0.020 | 0.063 | 0.748 | 1336/49 | 136/136 |
|  | Food Secure | 1 | No | 0.016 | 0.014 | 0.262 | 128/185 | 128/128 |
|  | Food Insecure | 1 | No | 0.017 | 0.065 | 0.793 | 136/49 | 136/136 |
|  | Food Secure | 5 | Yes | 0.002 | 0.015 | 0.888 | 80/124 | 124/124 |
|  | Food Insecure | 5 | Yes | 0.004 | 0.082 | 0.960 | 41/20 | 41/41 |
|  | Food Secure | 1 | Yes | 0.010 | 0.016 | 0.516 | 115/173 | 115/115 |
|  | Food Insecure | 1 | Yes | 0.018 | 0.070 | 0.796 | 92/49 | 92/92 |
